# A boost with SARS-CoV-2 BNT162b2 mRNA vaccine elicits strong humoral responses independently of the interval between the first two doses

**DOI:** 10.1101/2022.04.18.22273967

**Authors:** Alexandra Tauzin, Shang Yu Gong, Mark M. Painter, Rishi R. Goel, Debashree Chatterjee, Guillaume Beaudoin-Bussières, Lorie Marchitto, Marianne Boutin, Annemarie Laumaea, James Okeny, Gabrielle Gendron-Lepage, Catherine Bourassa, Halima Medjahed, Guillaume Goyette, Justine C. Williams, Yuxia Bo, Laurie Gokool, Chantal Morrisseau, Pascale Arlotto, Renée Bazin, Judith Fafard, Cécile Tremblay, Daniel E. Kaufmann, Gaston De Serres, Marceline Côté, Ralf Duerr, Valérie Martel-Laferrière, Allison R. Greenplate, E. John Wherry, Andrés Finzi

## Abstract

Due to the recrudescence of SARS-CoV-2 infections worldwide, mainly caused by Omicron BA.1 and BA.2 variants of concern, several jurisdictions are administering a mRNA vaccine boost. Here, we analyzed humoral responses induced after the second and third doses of mRNA vaccine in naïve and previously-infected donors who received their second dose with an extended 16-week interval. We observed that the extended interval elicited robust humoral responses against VOCs, but this response was significantly diminished 4 months after the second dose. Administering a boost to these individuals brought back the humoral responses to the same levels obtained after the extended second dose. Interestingly, we observed that administering a boost to individuals that initially received a short 3-4 weeks regimen elicited humoral responses similar to those elicited in the long interval regimen. Nevertheless, humoral responses elicited by the boost in naïve individuals did not reach those present in previously-infected vaccinated individuals.

## INTRODUCTION

Two years after the coronavirus disease 2019 (COVID-19) was declared pandemic by the WHO, the severe acute respiratory syndrome coronavirus 2 (SARS-CoV-2) continues to circulate worldwide and has evolved in several variants. The variants of concern (VOCs), defined as variants with increased transmissibility, virulence and/or against which vaccines and monoclonal antibody treatments are less effective (WHO, 2022a), are now the main source of concern about the evolving pandemic. Currently, the Delta and Omicron variants are the main circulating VOCs. The Delta (B.1.617.2) variant was declared as a VOC on May 2021 and the Omicron (B.1.1.529) variant in November 2021 (Choi and Smith, 2021; WHO, 2022a). Delta became the dominant strain in the summer/autumn of 2021. Omicron is divided into several sub-lineages: BA.1 (the main, named Omicron hereafter), BA.1.1, BA.2 and BA.3 (Kumar et al., 2022; Viana et al., 2022). Due to its relatively high number of mutations, notably in the Spike (S) glycoprotein, Omicron is more resistant to humoral responses elicited by vaccination or natural infection. This phenotype, in combination with a higher transmissibility rate compared to Delta, likely explains why it became the dominant strain worldwide by January 2022 (Chen et al., 2022; Dhar et al., 2021).

Vaccination campaigns began over a year ago and, in several parts of the world, public health authorities are administering a third dose of vaccine (boost). Vaccine scarcity at the beginning of the vaccination campaign led some public health authorities to increase the interval between the first two doses, notably in the province of Quebec, Canada, where this interval was delayed to 16 weeks instead of 3-4 weeks. Several studies have now shown that this strategy leads to improved humoral, T and B cell responses after the second dose in comparison to the short vaccine regimen, in particular against VOCs including Delta and Omicron variants (Chatterjee et al., 2021; Nayrac et al., 2021; Payne et al., 2021; Tauzin et al., 2022a).

A vaccine boost is now recommended in several jurisdictions worldwide in response to the Omicron wave (Ferdinands, 2022). Recent studies have shown that this boost, following the 3-4 weeks dose interval regimen, strongly improves humoral responses against VOCs, for which poor responses were observed after the second dose (Doria-Rose et al., 2021; Gruell et al., 2022; Nemet et al., 2022; Schmidt et al., 2022). Here, we analyzed humoral responses elicited after the second and the third dose of mRNA vaccine in a cohort of SARS-CoV-2 naïve and previously infected donors who received their first two doses of Pfizer BioNTech mRNA vaccine with a 16-weeks interval and compared to individuals receiving a short interval.

## RESULTS

We analyzed humoral responses induced after the second and the third doses of BNT162b2 mRNA vaccine in a cohort of donors who received their first two doses with an extended interval of 16 weeks (median [range]: 111 days [76–134 days]). These donors received their third dose around seven months after the second dose (median [range]: 219 days [167-235 days]). The cohort included 20 SARS-CoV-2 naïve and 11 previously infected (PI) individuals who tested SARS-CoV-2 positive by nasopharyngeal swab PCR around 10 months before their first dose (median [range]: 300 days [247-321 days]). Blood samples were analyzed three weeks (V3, median [range]: 21 days [13–42 days]) and four months (V4, median [range]: 112 days [90–156 days]) after the second dose and four weeks (V5, median [range]: 27 days [19–38 days]) after the third dose of mRNA vaccine. Basic demographic characteristics of the cohorts and detailed vaccination time points are summarized in Table 1 and Figure 1A.

**Table 1.** Characteristics of the vaccinated SARS-CoV-2 cohorts.

|  |  | SARS-CoV-2 Naïve | SARS-CoV-2 Previously infected |
| --- | --- | --- | --- |
| **Number** |  | 20 | 11 |
| **Age** |  | 52 (33-64) | 48 (39-65) |
| **Gender** | **Male (n)** | 8 | 8 |
|  | **Female (n)** | 12 | 3 |
| **Days between symptom onset and the 1<sup>st</sup> dose <sup>a</sup>** |  | N/A | 300 (247-321) |
| **Days between the 1<sup>st</sup> and 2<sup>nd</sup> dose <sup>a</sup>** |  | 111 (76-120) | 110 (90-134) |
| **Days between the 2<sup>nd</sup> and 3<sup>rd</sup> dose <sup>a</sup>** |  | 219 (167-230) | 219 (187-235) |
| **Days between the 2<sup>nd</sup> dose and V3 <sup>a</sup>** |  | 21 (17-34) | 22 (13-42) |
| **Days between the 2<sup>nd</sup> dose and V4 <sup>a</sup>** |  | 112 (96-156) | 113 (90-127) |
| **Days between the 3<sup>rd</sup> dose and V5 <sup>a</sup>** |  | 27 (20-38) | 27 (19-37) |

<sup>a</sup> Values displayed are medians, with ranges in parentheses.

**Figure 1.**
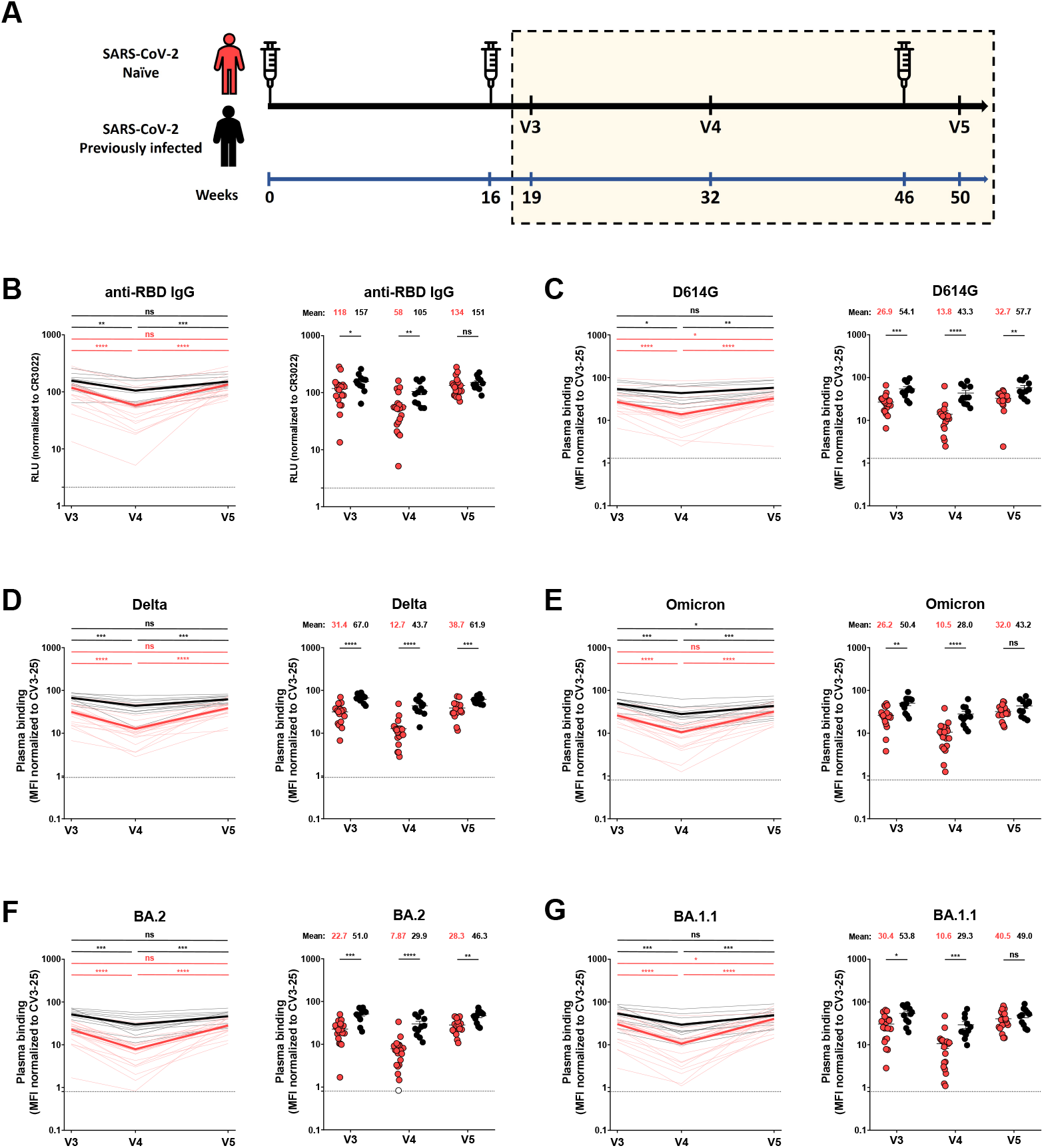
RBD-specific IgG and recognition of SARS-CoV-2 Spike variants by vaccine-elicited antibodies in SARS-CoV-2 naïve and previously-infected individuals after the second and the third dose of mRNA vaccine. (**A**) SARS-CoV-2 vaccine cohort design. The yellow box represents the period under study. (**B**) Indirect ELISA was performed by incubating plasma samples from naïve and PI donors collected at V3, V4 and V5 with recombinant SARS-CoV-2 RBD protein. Anti-RBD Ab binding was detected using HRP-conjugated anti-human IgG. Relative light unit (RLU) values obtained with BSA (negative control) were subtracted and further normalized to the signal obtained with the anti-RBD CR3022 mAb present in each plate. (**C-G**) 293T cells were transfected with the indicated full-length S from different SARS-CoV-2 variants S and stained with the CV3-25 Ab or with plasma from naïve or PI donors collected at V3, V4 and V5 and analyzed by flow cytometry. The values represent the median fluorescence intensities (MFI) normalized by CV3-25 Ab binding. (**B-G**) (**Left panels**) Each curve represents the values obtained with the plasma of one donor at every time point. Mean of each group is represented by a bold line. (**Right panels**) Plasma samples were grouped in different time points (V3, V4 and V5). Naïve and PI donors are represented by red and black points respectively. Undetectable measures are represented as white symbols, and limits of detection are plotted. Error bars indicate means ± SEM. (* p < 0.05; ** p < 0.01; *** p < 0.001; **** p < 0.0001; ns, non-significant). For naïve donors, n (number of individuals) = 20 and for previously infected donors n=11.

### Anti-RBD IgG levels of vaccine-elicited antibodies

We first measured the level of anti-receptor-binding domain (RBD) IgG induced after the second and the third doses of mRNA vaccine by ELISA assay (Anand et al., 2021; Beaudoin-Bussières et al., 2020; Prévost et al., 2020; Tauzin et al., 2021). Three weeks after the second dose of mRNA vaccine (V3), both naïve and PI individuals presented high levels of anti-RBD IgG (Figure 1B). Four months after the second dose (V4), the level of antibodies (Abs) decreased for both groups reaching significantly lower levels for naïve individuals, in agreement with previous observations (Tauzin et al., 2022a). The third dose (V5) led to an increase of anti-RBD IgG level, similar in both groups, that reached the same levels than after the second dose (V3) (Figure 1B).

### Recognition of SARS-CoV-2 Spike variants by plasma from vaccinated individuals

We evaluated the ability of plasma IgG to recognize SARS-CoV-2 full-length S variants after the second and the third dose of vaccine (Figure 1 C-G). After the second dose, plasma from naïve donors recognized the D614G S less efficiently than plasma from PI individuals (Figure 1C). Four months after the second dose (V4), we observed a decreased recognition for both groups, but more pronounced in the naïve group. The third dose (V5) increased D614G S recognition by the naïve group, reaching levels similar to those achieved after the second dose (V3) (Figure 1C). However, even after the boost, the level of recognition in naïve donors did not reach the same level as in the PI group.

The original Wuhan strain was used to develop mRNA SARS-CoV-2 vaccines. Numerous mutations, particularly in the S glycoprotein, reduced the ability of vaccine-induced Abs to recognize currently circulating strains. We tested the S recognition of several VOCs in circulation after mRNA vaccination (Figure 1D-G, S2C-E). For all the VOCs S tested, a similar pattern of response than for D614G S was observed. Except for Omicron and BA.1.1 S at V5, plasma from PI donors more efficiently recognized S variants than naïve donors at all time points. Again, booster-elicited antibodies able to recognize the different S glycoproteins reached levels similar as those obtained after the second dose (Figure 1D-G). Comparable responses were observed when we measured the capacity of plasma to recognize the S2 subunit (Figure S1A).

We also evaluated whether the booster impacted the capacity of plasma to recognize the S glycoprotein of the endemic HKU1 human *Betacoronaviruses* (Figure S1B). We did not observe major changes in recognition after the second and third doses. However, for all time points plasma from PI donors always recognized better the HCoV-HKU1 S than the naïve group. This suggests that natural infection elicits more cross-reactive antibodies.

### Functional activities of vaccine-elicited antibodies

We evaluated functional activities of vaccine-elicited Abs after the second and third doses (Figure 2). We measured Fc-effector functions using a well-described antibody-dependent cellular cytotoxicity (ADCC) assay (Anand et al., 2021; Beaudoin-Bussières et al., 2020, 2021; Ullah et al., 2021). Plasma from PI individuals presented significantly higher ADCC activity after the second dose (V3 and V4) with naïve individuals reaching similar levels after the third dose (V5) (Figure 2A). We noted that while ADCC remained relatively stable over time for PI individuals, it significantly decreased 4 months after the second dose for naïve individuals (V4). A similar pattern of responses was observed for the neutralizing activity against pseudoviruses carrying the D614G S (Figure 2B). At the three time points, the neutralizing activity was better in the PI group compared to naïve group. The level of neutralizing Abs remained stable in PI individuals whereas it significantly decreased in naïve donors at V4 but was significantly increased by the boost.

**Figure 2.**
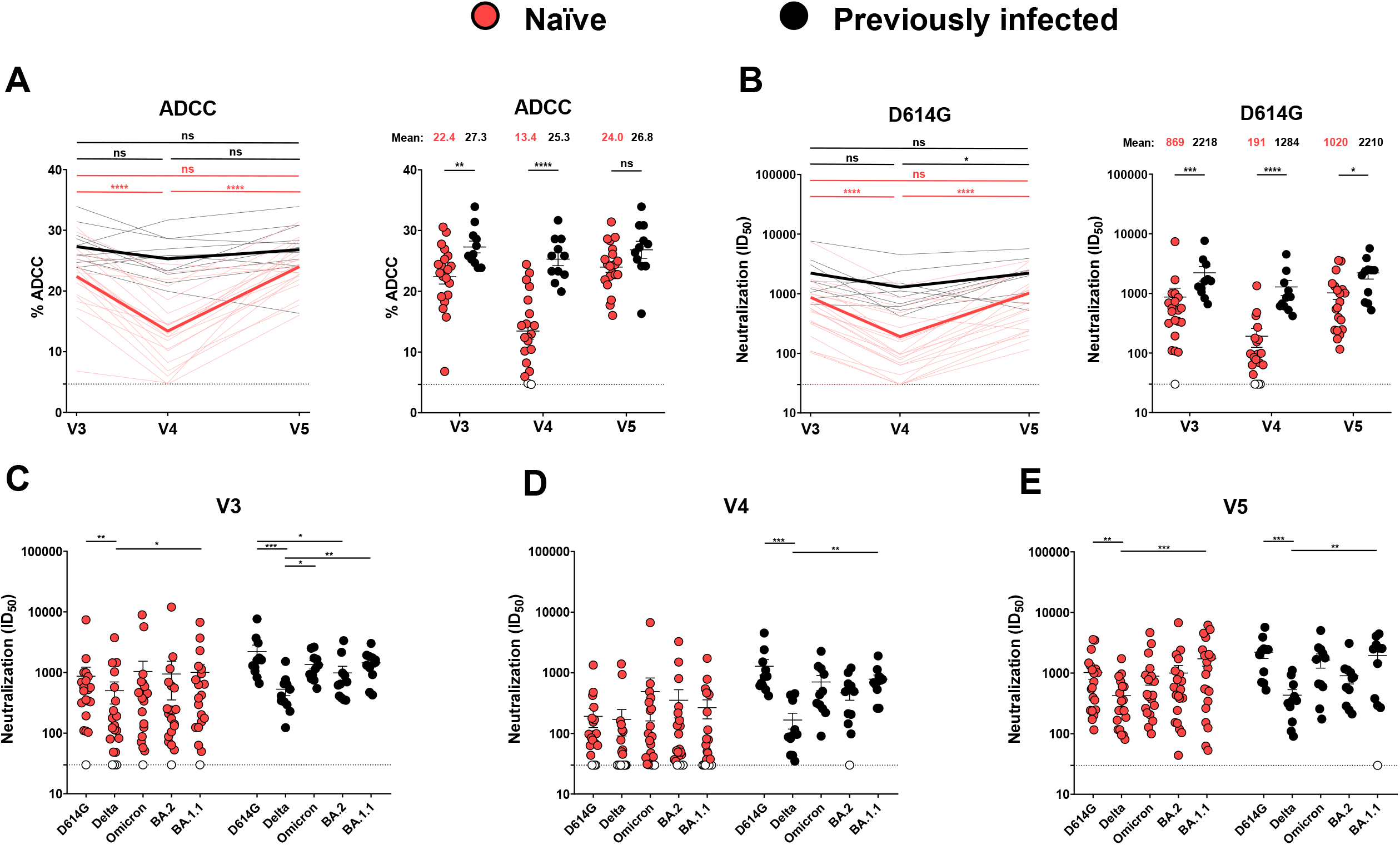
Fc-effector functions and neutralization activities induced by mRNA vaccination in SARS-CoV-2 naïve and previously-infected individuals. (**A**) CEM.NKr parental cells were mixed at a 1:1 ratio with CEM.NKr-Spike cells and were used as target cells. PBMCs from uninfected donors were used as effector cells in a FACS-based ADCC assay. (**B-E**) Neutralizing activity was measured by incubating pseudoviruses bearing SARS-CoV-2 S glycoproteins, with serial dilutions of plasma for 1 h at 37°C before infecting 293T-ACE2 cells. Neutralization half maximal inhibitory serum dilution (ID_50_) values were determined using a normalized non-linear regression using GraphPad Prism software. (**A-B**) (**Left panels**) Each curve represents the values obtained with the plasma of one donor at every time point. Mean of each group is represented by a bold line. (**Right panels**) Plasma samples were grouped in different time points (V3, V4 and V5). (**C-E**) Neutralization activities against several SARS-CoV-2 variants S were analyzed at the different time points (V3 (**C**), V4 (**D**) and V5 (**E**)). Naïve and PI donors are represented by red and black points respectively. Undetectable measures are represented as white symbols, and limits of detection are plotted. Error bars indicate means ± SEM. (* p < 0.05; ** P < 0.01; *** p < 0.001; **** p < 0.0001; ns, non-significant). For naïve donors, n=20 and for previously infected donors n=11.

When looking at the neutralizing activity against VOCs, we observed that plasma from PI group more efficiently neutralized all pseudoviruses than the naïve group after the second dose (Figure 2C-D, S2). Interestingly, this difference disappeared after the boost (V5) (Figure S2).

### Integrated analysis of vaccine responses elicited by the second and third doses

We evaluated the network of pairwise correlations among all studied immune variables on 11 randomly selected naïve donors and the 11 PI individuals. For SARS-CoV-2 naïve individuals (Figure 3A), we observed a dense network of positive correlations after the second dose (V3) involving all immune variables tested, except for HCoV-HKU1 S binding. Four months after the second dose, this network became less dense, and the third dose did not substantially alter the network of correlations except for improved correlations of neutralization responses against Omicron and BA.1.1 with other anti-SARS-CoV-2 immune responses. Interestingly, in PI individuals, the integrated network was less dense after the second dose compared to naïve donors (Figure 3B), suggesting a less focused immune response possibly due to the heterogenous immune stimulations by natural infection and vaccination. Whereas the network became slightly denser among the S binding responses four months after the second dose, it remained sparsely connected overall without major changes after the third dose of the mRNA vaccine.

**Figure 3.**
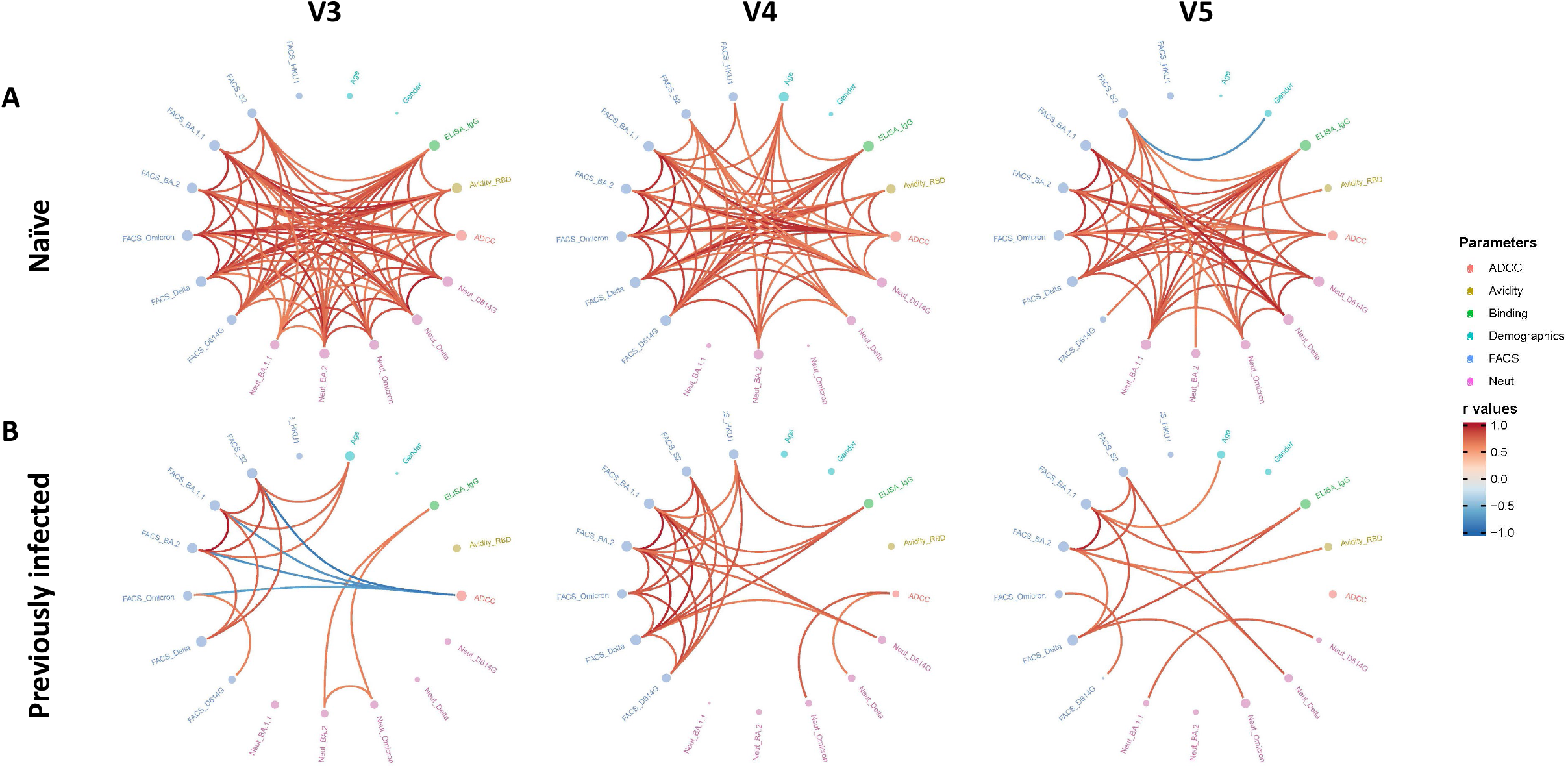
Mesh correlations of humoral response variables after the second and third dose of the mRNA vaccine. Edge bundling correlation plots where red and blue edges represent positive and negative correlations between connected variables, respectively. Only significant correlations (p < 0.05, Spearman rank test) are displayed. Nodes are color coded based on the grouping of variables according to the legend. Node size corresponds to the degree of relatedness of correlations. Edge bundling plots are shown for correlation analyses using six different datasets, i.e., SARS-CoV-2 naïve (**A**) or previously infected (**B**) individuals at V3, V4 and V5 respectively. For naïve and previously infected donors: n=11.

### Evolution of anti-RBD avidity induced after a short or a long interval between mRNA vaccine doses

We and others previously described that an extended interval between the first two doses of mRNA vaccine led to better humoral and cellular responses than the 3-4 weeks standard regimen, especially against VOCs (Nayrac et al., 2021; Payne et al., 2021; Tauzin et al., 2022a). However, whether the humoral advantages observed with the long interval persist after the boost remains unclear. To address this important question, we measured longitudinally the level of anti-RBD IgG in cohorts of naïve and PI individuals that received their first two doses with the standard (short interval, SI) or the extended regimen (long interval, LI). Basic demographic characteristics of the cohorts and detailed vaccination time points are summarized in Table 2 and Figure 4A. Collection time points did not perfectly match between the two cohorts rendering perilous a side-by-side comparison using assays that only measure antibody quantities rather than their quality. We therefore decided to measure the avidity for the RBD of the induced IgG by a previously described assay (Björkman et al., 1999; Fialová et al., 2017; Tauzin et al., 2022b). This assay consists of parallel ELISAs with washing buffer having or not a chaotropic agent (8M urea). The RBD-avidity index therefore becomes a surrogate of antibody maturation since only Abs with the highest avidity remain attached to the RBD after 8M urea washing (Figure S3).

**Table 2.** Characteristics of the longitudinal SARS-CoV-2 cohorts vaccinated with a short or a long interval.

|  |  | Short interval |  | Long interval |  |
| --- | --- | --- | --- | --- | --- |
|  |  | SARS-CoV-2 Naïve | SARS-CoV-2 Previously infected | SARS-CoV-2 Naïve | SARS-CoV-2 Previously infected |
| Number |  | 45 | 16 | 30 | 15 |
| Age |  | 35 (22-67) | 35 (23-59) | 51 (21-64) | 47 (29-65) |
| Gender | Male (n) | 21 | 10 | 12 | 10 |
|  | Female (n) | 24 | 6 | 18 | 5 |
| Days between symptom onset and the 1<sup>st</sup> dose <sup>a</sup> |  | N/A | 102 (50-275) | N/A | 274 (166-321) |
| Days between the 1<sup>st</sup> and 2<sup>nd</sup> dose <sup>a</sup> |  | 21 (21-35) | 21 (21-30) | 111 (76-120) | 110 (90-134) |
| Days between the 2<sup>nd</sup> and 3<sup>rd</sup> dose <sup>a</sup> |  | 258 (183-342) | 276 (244-308) | 219 (167-230) | 219 (187-235) |

<sup>a</sup> Values displayed are medians, with ranges in parentheses.

**Figure 4.**
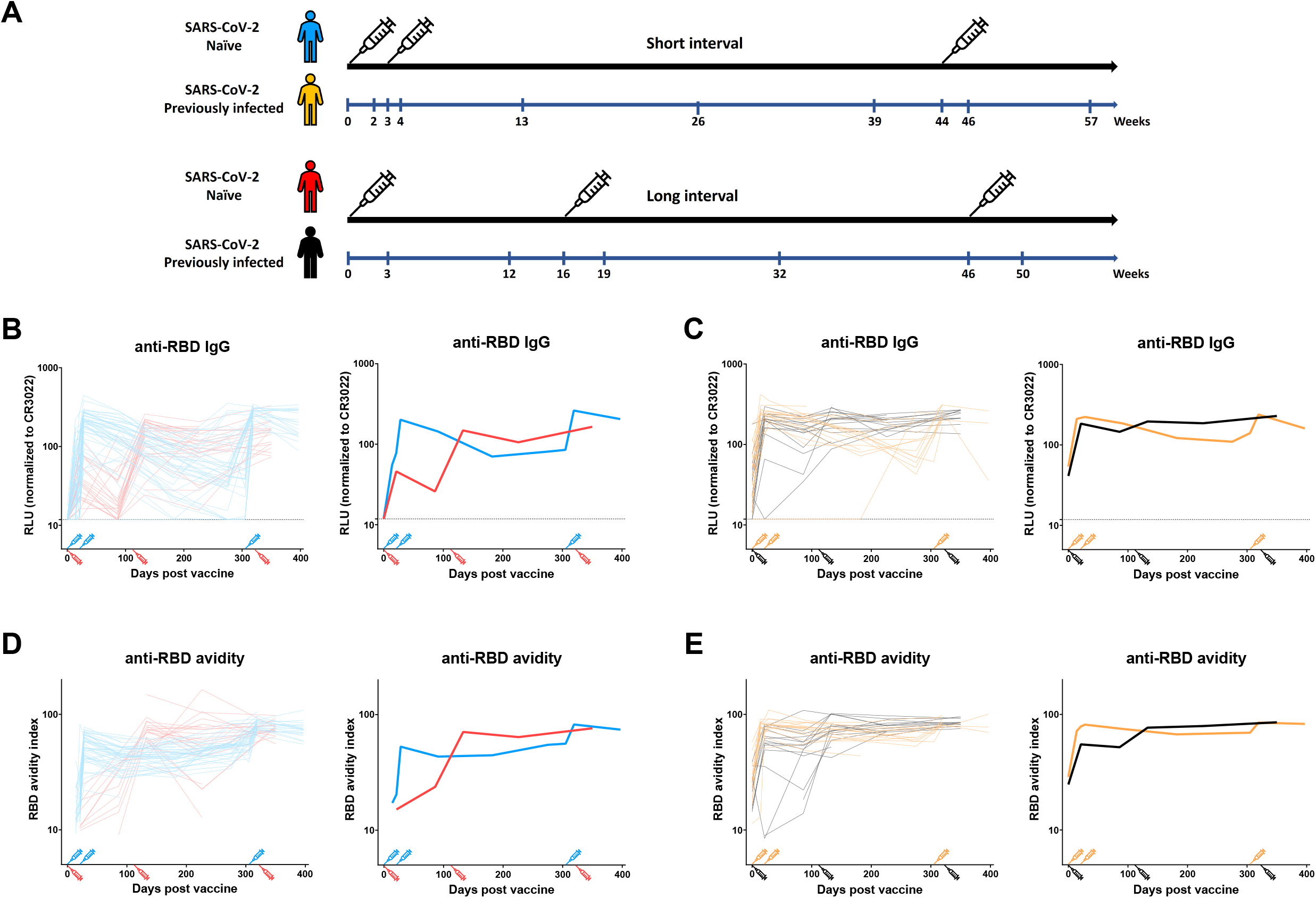
Evolution of the RBD-specific IgG and associated anti-RBD avidity in SARS-CoV-2 naïve and previously-infected individuals vaccinated with a short or a long interval. (**A**) SARS-CoV-2 vaccine cohorts design. (**B-E**) Indirect ELISA were performed by incubating plasma samples from naïve (**B, D**) and PI (**C, E**) donors vaccinated with a SI or a LI with recombinant SARS-CoV-2 RBD protein. The plasmas were collected at different time point from prior vaccination to after the third dose of mRNA vaccine. Anti-RBD Ab binding was detected using HRP-conjugated anti-human IgG. (**B-C**) Relative light unit (RLU) values obtained were normalized to the signal obtained with the anti-RBD CR3022 mAb present in each plate. (**D-E)** The RBD avidity index corresponded to the value obtained with the stringent (8M urea) ELISA divided by that obtained without urea. (**B-E**) (**Left panels**) Each curve represents the values obtained with the plasma of one donor at every time point. (**Right panels**) The bold line represents the mean of each group. Naïve and PI donors vaccinated with the SI are represented by blue and yellow lines respectively and naïve and PI donors vaccinated with the LI are represented by red and black lines respectively. The time of vaccine dose injections is indicated by an associated colour syringe. Limits of detection are plotted. For naïve donors, n= 45 for the SI and n=30 for the LI and for previously infected donors n=16 for the SI and n=15 for the LI.

Anti-RBD antibodies reached their peak level faster with the SI compared to the LI in naïve individuals (Figure 4B). While antibody levels rapidly decreased after the second dose in the SI regimen, the decay in the LI group was slower. In both groups, a booster elicited the highest levels of antibodies. Consistent with the proposed use of the RBD-avidity index as a surrogate for antibody maturation, the kinetics differed from those of the regular ELISA that only measure total levels of anti-RBD Abs. For example, 12 weeks after the first dose in the LI regimen, there was a significant decline in anti-RBD IgG levels but the affinity of anti-RBD Abs likely improved as shown by an increase in their avidity (Figure 4B and D), consistent with recent results (Tauzin et al., 2022b). We previously reported that several humoral responses, including RBD-avidity was lower in individuals receiving a SI (Tauzin et al., CHM 2022). However, whether this difference remained after the boost was unknown. Here we report that the boost brought the RBD-avidity index to the same level, independent of the vaccine regimen, suggesting that the boost is required to further improve antibody responses in naïve individuals that received the SI regimen (Figure 4D).

For the PI groups, the SI and LI led rapidly to high levels of IgG (Figure 4C). These levels slightly decreased over time, however they remained more stable in the LI group, likely due to the delayed dose. After the boost, we observed similar levels of IgG in both groups. Regarding the avidity, the SI rapidly led to IgG with strong avidity which remained stable over time with only a minor effect induced by the boost (Figure 4E). For the LI group, the first dose increased the avidity, but to a lower extend than in the SI group. The second dose boosted the avidity to the same level than in the SI group. As observed in the SI group, the boost did not improve the avidity, indicating that individuals that developed hybrid immunity due to natural infection already had a higher antibody avidity than naïve vaccinated individuals.

## DISCUSSION

Currently, a large part of the world population has received two or three doses of SARS-CoV-2 mRNA vaccines (WHO, 2022b). These vaccines were based on the ancestral Wuhan strain. One of the major challenges of the pandemic is the frequent emergence of variants that are more resistant to vaccine-elicited humoral responses, thus fueling new waves. In this study, we found that individuals who received their first two doses of mRNA vaccine with a 16-week extended interval had strong humoral responses against VOCs induced after the second dose. These responses significantly decreased 4 months later but came back to peak levels after the boost.

Many studies have shown that an extended interval between doses elicits humoral responses that outperform those elicited by a short interval in SARS-CoV-2 naïve individuals. Accordingly, we observed a network of correlation less dense after the second dose in individuals vaccinated with a SI in comparison to a LI (Figure 3A and S4). However, the third dose of mRNA vaccine, administered several months after the second dose in the SI regimen, strongly improved the humoral responses against VOCs (Figure S5), and the network of correlations became denser, as observed after the LI regimen (Figure 3A and S4), emphasising the importance of administering the boost.

Antibody maturation is a complex process that takes place in the germinal center (Young and Brink, 2021). This process is important for vaccine efficacy since it allows the immune system to generate antibodies with greater potency for viral neutralization and Fc-effector functions. We recently developed a high-throughput assay that could be used as a surrogate for antibody maturation (Tauzin et al., 2022b). Using this assay, we observed that the avidity of anti-RBD Abs was significantly higher in the LI after the second dose compared to the SI, suggesting that increasing the time between exposure to the antigen led to a better maturation of the B cells and so to Abs with higher avidity (Tauzin et al., 2022a). Here we report that a boost allows the SI group to elicit Abs with the same avidity as in the LI group. Since the RBD is highly immunogenic, it is possible that humoral responses against this domain compromise responses against less immunogenic domains of the Spike. Thus, in future studies it would be interesting to measure the evolution of the avidity against other Spike domains.

Vaccinated PI individuals presented a better avidity than naïve individuals at all time points. These results are consistent with previous observations indicating that hybrid immunity led to broad and stronger humoral responses, but the mechanisms remain unclear (Andreano et al., 2021; Crotty, 2021; Goel et al., 2021). In correlation networks after the second and third doses of mRNA vaccine, we observed strikingly different profiles of correlations in naïve compared to PI donors suggesting that infection primes the immune system in a different way than vaccination does. Whether this is linked to the immune stimulation with all components of the entire virus or the transmission mode of SARS-CoV-2 which infects host by the mucosa, therefore activating resident immune cells, remains poorly understood.

While vaccination confers good protection against severe COVID-19, it is less efficient against viral transmission. Thus, breakthrough infection in vaccinated individuals appears frequently. It was recently shown that breakthrough infection in vaccinated individuals induced strong neutralizing Abs efficient against VOCs, including Omicron (Miyamoto et al., 2022). Interestingly, a LI between vaccination and breakthrough infection also induced better humoral responses against VOCs than a SI, as observed for vaccination. It is possible that breakthrough infection in fully vaccinated individuals led to hybrid immunity with humoral responses as strong as those observed in infected-then-vaccinated individuals. Moreover, it will be interesting to see the impact of breakthrough infection on the avidity of the Abs.

The third dose of SARS-CoV-2 mRNA vaccine led to high levels of humoral responses against VOCs, irrespective of the interval between the two first doses. We do not yet know about the durability of this immunity but epidemiological studies have shown a decline in vaccine effectiveness against Omicron within a few months of the third dose (Andrews et al., 2022). We observed a rapid decrease of Ab levels after the first and second dose with both intervals and to a greater extent in SARS-CoV-2 naïve donors. However, it is possible that while humoral responses rapidly decreased after vaccination, cellular responses remain stable. Monitoring the evolution of humoral and cellular responses after the third dose of vaccine, and in particular in the numerous individuals who had Omicron or BA.2 breakthrough infection after their third dose, will be necessary to determine the need for additional boosts, the best interval time between dose injections, and the populations to be targeted (general population or only population at risk).

## Data Availability

All data produced in the present work are contained in the manuscript

## ACKNOWLEDGMENTS

The authors are grateful to the donors who participated in this study. The authors thank the CRCHUM BSL3 and Flow Cytometry Platforms for technical assistance. We thank Dr. Stefan Pöhlmann (Georg-August University, Germany) for the plasmid coding for SARS-CoV-2 S glycoproteins and Dr. M. Gordon Joyce (U.S. MHRP) for the monoclonal antibody CR3022. This work was supported by le Ministère de l’Économie et de l’Innovation du Québec, Programme de soutien aux organismes de recherche et d’innovation to A.F. and by the Fondation du CHUM. This work was also supported by a CIHR foundation grant #352417, by a CIHR operating Pandemic and Health Emergencies Research grant #177958, a CIHR stream 1 and 2 for SARS-CoV-2 Variant Research to A.F., and by an Exceptional Fund COVID-19 from the Canada Foundation for Innovation (CFI) #41027 to A.F. and D.E.K. Work on variants presented was also supported by the Sentinelle COVID Quebec network led by the LSPQ in collaboration with Fonds de Recherche du Québec Santé (FRQS) to A.F. This work was also partially supported by a CIHR COVID-19 rapid response grant (OV3 170632) and CIHR stream 1 SARS-CoV-2 Variant Research to M.C. A.F. is the recipient of Canada Research Chair on Retroviral Entry no. RCHS0235 950-232424. M.C is a Tier II Canada Research Chair in Molecular Virology and Antiviral Therapeutics. V.M.L. is supported by a FRQS Junior 1 salary award. D.E.K is a FRQS Merit Research Scholar. G.B.B. is the recipient of an FRQS PhD fellowship. A.L. was supported by MITACS Accélération postdoctoral fellowships. This work was also supported by NIH grants AI108545, AI155577, AI149680, and U19AI082630 (to E.J.W.), the University of Pennsylvania Perelman School of Medicine COVID Fund (to R.R.G. and E.J.W.); the University of Pennsylvania Perelman School of Medicine 21st Century Scholar Fund (to R.R.G.); and the Paul and Daisy Soros Fellowship for New Americans (to R.R.G). The funders had no role in study design, data collection and analysis, decision to publish, or preparation of the manuscript. We declare no competing interests.

## AUTHOR CONTRIBUTIONS

A.T. and A.F. conceived the study. A.T., S.Y.G., D.C., G.B.B., L.M., M.B., A.L., C.B., G.G.L, Y.B., and A.F. performed, analyzed, and interpreted the experiments. A.T. and R.D. performed statistical analysis. S.Y.G., M.M.P., R.R.G, G.B.B., A.L., J.O., G.G.L., H.M., G.G., Y.B., M.C, A.R.G., E.J.W. and A.F. contributed unique reagents. J.C.W., L.G., C.M., P.A., C.T., D.E.K., and V.M.-L. collected and provided clinical samples. G.D.S., J.F., D.E.K., E.J.W., and R.B. provided scientific input related to VOC and vaccine efficacy. A.T. and A.F. wrote the manuscript with inputs from others. Every author has read, edited, and approved the final manuscript.

## STAR METHODS

## RESOURCE AVAILABILITY

### Lead contact

Further information and requests for resources and reagents should be directed to and will be fulfilled by the lead contact, Andrés Finzi.

### Materials availability

All unique reagents generated during this study are available from the Lead contact without restriction.

### Data and code availability

- All data reported in this paper will be shared by the lead contact upon request.
- This paper does not report original code.
- Any additional information required to reanalyze the data reported in this paper is available from the lead contact upon request.

## EXPERIMENTAL MODEL AND SUBJECT DETAILS

### Ethics Statement

All work was conducted in accordance with the Declaration of Helsinki in terms of informed consent and approval by an appropriate institutional board. Blood samples were obtained from donors who consented to participate in this research project at CHUM (19.381) and University of Pennsylvania (University of Pennsylvania Institutional Review Board, IRB no. 845061). Plasmas were isolated by centrifugation and Ficoll gradient, and samples stored at -80°C until use.

### Human subjects

The study was conducted in 20 SARS-CoV-2 naïve individuals (8 males and 12 females; age range: 33-64 years) and 11 SARS-CoV-2 previously-infected individuals (8 males and 3 females; age range: 39-65 years). All this information is summarized in table 1. For the comparison between the SI and LI, the study was conducted in 45 SARS-CoV-2 naïve individuals (21 males and 24 females ; age range : 22-67 years) and 16 SARS-CoV-2 previously-infected individuals (10 males and 6 females ; age range : 23-59 years) for the SI and in 30 SARS-CoV-2 naïve individuals (12 males and 18 females ; age range : 21-64 years) and 15 SARS-CoV-2 previously-infected individuals (10 males and 5 females ; age range : 29-65 years) for the LI. All this information is summarized in table 2. No specific criteria such as number of patients (sample size), gender, clinical or demographic were used for inclusion, beyond PCR confirmed SARS-CoV-2 infection in adults and no detection of Abs recognizing the N protein for naïve donors.

### Plasma and antibodies

Plasma from SARS-CoV-2 naïve and PI donors were collected, heat-inactivated for 1 hour at 56°C and stored at -80°C until ready to use in subsequent experiments. Plasma from uninfected donors collected before the pandemic were used as negative controls and used to calculate the seropositivity threshold in our ELISA, ADCC and flow cytometry assays (see below). The RBD-specific monoclonal antibody CR3022 was used as a positive control in ELISA assays, and the CV3-25 antibody in flow cytometry assays and were previously described (Anand et al., 2020; Beaudoin-Bussières et al., 2020; Jennewein et al., 2021; Meulen et al., 2006; Prévost et al., 2020). Horseradish peroxidase (HRP)-conjugated Abs able to detect the Fc region of human IgG (Invitrogen) was used as secondary Abs to detect Ab binding in ELISA experiments. Alexa Fluor-647-conjugated goat anti-human Abs able to detect all Ig isotypes (anti-human IgM+IgG+IgA; Jackson ImmunoResearch Laboratories) were used as secondary Ab to detect plasma binding in flow cytometry experiments.

### Cell lines

293T human embryonic kidney cells (obtained from ATCC) were maintained at 37°C under 5% CO_2_ in Dulbecco’s modified Eagle’s medium (DMEM) (Wisent) containing 5% fetal bovine serum (FBS) (VWR) and 100 μg/ml of penicillin-streptomycin (Wisent). CEM.NKr CCR5+ cells (NIH AIDS reagent program) were maintained at 37°C under 5% CO_2_ in Roswell Park Memorial Institute (RPMI) 1640 medium (Gibco) containing 10% FBS and 100 μg/ml of penicillin-streptomycin. 293T-ACE2 cell line was previously reported (Prévost et al., 2020). CEM.NKr CCR5+ cells stably expressing the SARS-CoV-2 S glycoproteins were previously reported (Anand et al., 2021; Beaudoin-Bussières et al., 2021).

## METHOD DETAILS

### Plasmids

The plasmids encoding the SARS-CoV-2 S variants (D614G, Delta (B.1.617.2) and Omicron (B.1.1.529) and the S2 subunit were previously reported (Beaudoin-Bussières et al., 2020; Chatterjee et al., 2021; Gong et al., 2021; Tauzin et al., 2022a). The HCoV-HKU1 S was purchased from Sino Biological. The plasmids encoding the BA.1.1 and BA.2 S were generated by overlapping PCR for mutagenesis of a codon-optimized wild-type SARS-CoV-2 S gene (GeneArt, ThermoFisher) that was synthesized (Biobasic) and cloned in pCAGGS as a template. All constructs were verified by Sanger sequencing.

### Protein expression and purification

FreeStyle 293F cells (Invitrogen) were grown in FreeStyle 293F medium (Invitrogen) to a density of 1 × 10^6^ cells/mL at 37°C with 8% CO_2_ with regular agitation (150 rpm). Cells were transfected with a plasmid coding for SARS-CoV-2 S RBD (Beaudoin-Bussières et al., 2020) using ExpiFectamine 293 transfection reagent, as directed by the manufacturer (Invitrogen). One week later, cells were pelleted and discarded. Supernatants were filtered using a 0.22 µm filter (Thermo Fisher Scientific). The recombinant RBD proteins were purified by nickel affinity columns, as directed by the manufacturer (Invitrogen). The RBD preparations were dialyzed against phosphate-buffered saline (PBS) and stored in aliquots at -80°C until further use. To assess purity, recombinant proteins were loaded on SDS-PAGE gels and stained with Coomassie Blue.

### Enzyme-Linked Immunosorbent Assay (ELISA) and RBD avidity index

The SARS-CoV-2 RBD ELISA assay used was previously described (Beaudoin-Bussières et al., 2020; Prévost et al., 2020). Briefly, recombinant SARS-CoV-2 S RBD proteins (2.5 μg/ml), or bovine serum albumin (BSA) (2.5 μg/ml) as a negative control, were prepared in PBS and were adsorbed to plates (MaxiSorp Nunc) overnight at 4°C. Coated wells were subsequently blocked with blocking buffer (Tris-buffered saline [TBS] containing 0.1% Tween20 and 2% BSA) for 1h at room temperature. Wells were then washed four times with washing buffer (Tris-buffered saline [TBS] containing 0.1% Tween20). CR3022 mAb (50 ng/ml) or a 1/500 dilution of plasma were prepared in a diluted solution of blocking buffer (0.1 % BSA) and incubated with the RBD-coated wells for 90 minutes at room temperature. Plates were washed four times with washing buffer followed by incubation with secondary Abs (diluted in a diluted solution of blocking buffer (0.4% BSA)) for 1h at room temperature, followed by four washes. To calculate the RBD-avidity index, we performed in parallel a stringent ELISA, where the plates were washed with a chaotropic agent, 8M of urea, added of the washing buffer. This assay was previously described (Tauzin et al., 2022b). HRP enzyme activity was determined after the addition of a 1:1 mix of Western Lightning oxidizing and luminol reagents (Perkin Elmer Life Sciences). Light emission was measured with a LB942 TriStar luminometer (Berthold Technologies). Signal obtained with BSA was subtracted for each plasma and was then normalized to the signal obtained with CR3022 present in each plate. The seropositivity threshold was established using the following formula: mean of pre-pandemic SARS-CoV-2 negative plasma + (3 standard deviation of the mean of pre-pandemic SARS-CoV-2 negative plasma).

### Cell surface staining and flow cytometry analysis

293T cells were co-transfected with a GFP expressor (pIRES2-GFP, Clontech) in combination with plasmids encoding the full-length S of SARS-CoV-2 variants (D614G, Delta and Omicron, BA.1.1 and BA.2), the S2 subunit or the HCoV-HKU1 S. 48h post-transfection, S-expressing cells were stained with the CV3-25 Ab (Jennewein et al., 2021) or plasma (1/250 dilution). AlexaFluor-647-conjugated goat anti-human IgM+IgG+IgA Abs (1/800 dilution) were used as secondary Abs. The percentage of transfected cells (GFP+ cells) was determined by gating the living cell population based on viability dye staining (Aqua Vivid, Invitrogen). Samples were acquired on a LSRII cytometer (BD Biosciences) and data analysis was performed using FlowJo v10.7.1 (Tree Star). The seropositivity threshold was established using the following formula: mean of pre-pandemic SARS-CoV-2 negative plasma + (3 standard deviation of the mean of pre-pandemic SARS-CoV-2 negative plasma). The conformational-independent S2-targeting mAb CV3-25 was used to normalize S expression. CV3-25 was shown to effectively recognize all SARS-CoV-2 S variants (Chatterjee et al., 2021; Li et al., 2022).

### ADCC assay

This assay was previously described (Anand et al., 2021; Beaudoin-Bussières et al., 2021). For evaluation of anti-SARS-CoV-2 antibody-dependent cellular cytotoxicity (ADCC), parental CEM.NKr CCR5+ cells were mixed at a 1:1 ratio with CEM.NKr cells stably expressing a GFP-tagged full length SARS-CoV-2 S (CEM.NKr.SARS-CoV-2.Spike cells). These cells were stained for viability (AquaVivid; Thermo Fisher Scientific, Waltham, MA, USA) and cellular dyes (cell proliferation dye eFluor670; Thermo Fisher Scientific) to be used as target cells. Overnight rested PBMCs were stained with another cellular marker (cell proliferation dye eFluor450; Thermo Fisher Scientific) and used as effector cells. Stained target and effector cells were mixed at a ratio of 1:10 in 96-well V-bottom plates. Plasma (1/500 dilution) or monoclonal antibody CR3022 (1 µg/mL) were added to the appropriate wells. The plates were subsequently centrifuged for 1 min at 300g, and incubated at 37°C, 5% CO_2_ for 5 hours before being fixed in a 2% PBS-formaldehyde solution. ADCC activity was calculated using the formula: [(% of GFP+ cells in Targets plus Effectors) -(% of GFP+ cells in Targets plus Effectors plus plasma/antibody)]/(% of GFP+ cells in Targets) x 100 by gating on transduced live target cells. All samples were acquired on an LSRII cytometer (BD Biosciences) and data analysis was performed using FlowJo v10.7.1 (Tree Star). The specificity threshold was established using the following formula: mean of pre-pandemic SARS-CoV-2 negative plasma + (3 standard deviation of the mean of pre-pandemic SARS-CoV-2 negative plasma).

### Virus neutralization assay

To produce the pseudoviruses, 293T cells were transfected with the lentiviral vector pNL4.3 R-E-Luc (NIH AIDS Reagent Program) and a plasmid encoding for the indicated S glycoprotein (D614G, Delta and Omicron, BA.1.1 and BA.2) at a ratio of 10:1. Two days post-transfection, cell supernatants were harvested and stored at -80°C until use. For the neutralization assay, 293T-ACE2 target cells were seeded at a density of 1×10^4^ cells/well in 96-well luminometer-compatible tissue culture plates (Perkin Elmer) 24h before infection. Pseudoviral particles were incubated with several plasma dilutions (1/50; 1/250; 1/1250; 1/6250; 1/31250) for 1h at 37°C and were then added to the target cells followed by incubation for 48h at 37°C. Then, cells were lysed by the addition of 30 µL of passive lysis buffer (Promega) followed by one freeze-thaw cycle. An LB942 TriStar luminometer (Berthold Technologies) was used to measure the luciferase activity of each well after the addition of 100 µL of luciferin buffer (15mM MgSO_4_, 15mM KPO_4_ [pH 7.8], 1mM ATP, and 1mM dithiothreitol) and 50 µL of 1mM d-luciferin potassium salt (Prolume). The neutralization half-maximal inhibitory dilution (ID_50_) represents the plasma dilution to inhibit 50% of the infection of 293T-ACE2 cells by pseudoviruses.

## QUANTIFICATION AND STATISTICAL ANALYSIS

### Statistical analysis

Symbols represent biologically independent samples from SARS-CoV-2 naïve or SARS-CoV-2 PI individuals. Lines connect data from the same donor. Statistics were analyzed using GraphPad Prism version 8.0.1 (GraphPad, San Diego, CA). Every dataset was tested for statistical normality and this information was used to apply the appropriate (parametric or nonparametric) statistical test. Differences in responses for the same patient before and after vaccination were performed using Wilcoxon tests. Differences in responses between naïve and PI individuals at each time point (V3, V4 and V5) were measured by Mann-Whitney tests. Differences in responses against the different S for the same patient were measured by Friedman tests. P values < 0.05 were considered significant; significance values are indicated as *p < 0.05, **p < 0.01, ***p < 0.001, ****p < 0.0001. Spearman’s R correlation coefficient was applied for correlations. Statistical tests were two-sided and p < 0.05 was considered significant.

### Software scripts and visualization

Edge bundling graphs were generated in undirected mode in R and RStudio using ggraph, igraph, tidyverse, and RColorBrewer packages (R Core Team, 2014). Edges are only shown if p < 0.05, and nodes are sized according to the connecting edges’ r values. Nodes are color-coded according to groups of parameters.

---

**Figure S1:**
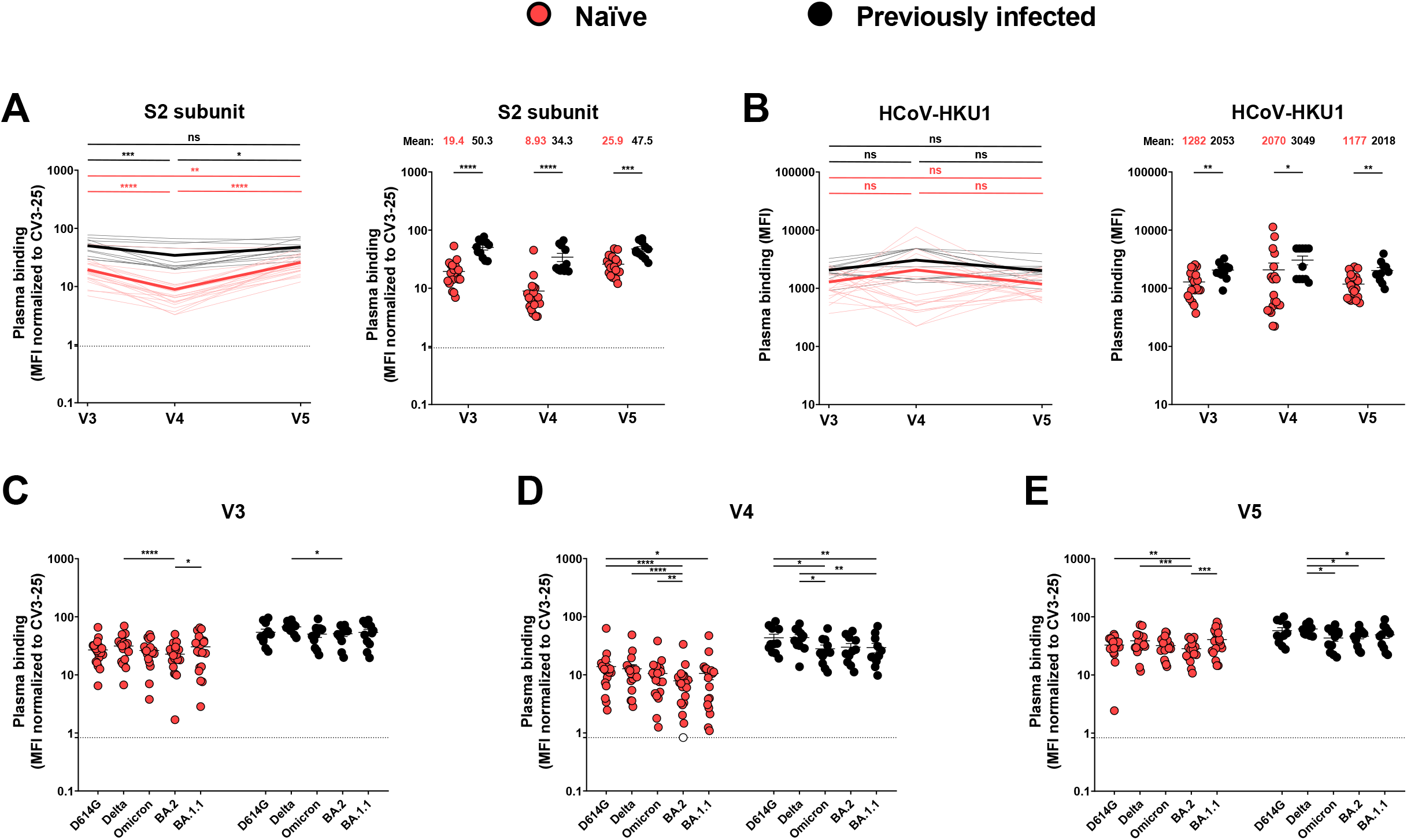
Recognition of SARS-CoV-2 Spike variants and HCoV-HKU1 S by plasma from naïve and PI donors, Related to Figure 1. 293T cells were transfected with the S2 subunit (**A**) or the indicated full-length S from different SARS-CoV-2 variants (**C-E**) or the HCoV-HKU1 S (**B**) and stained with the CV3-25 Ab or with plasma from naive or PI donors collected at V3, V4 and V5 and analyzed by flow cytometry. The values represent the MFI (**B**) or the MFI normalized by CV3-25 Ab binding (**A, C-E**). (**A-B**) Left panel: Each curve represents the values obtained with the plasma of one donor at every time point. Mean of each group is represented by a bold line. Right panel: Plasma samples were grouped in different time points (V3, V4 and V5). (**C-E**) Binding of plasma collected at V3 (**C**), V4 (**D**) and V5 (**E**). Naïve and PI donors are represented by red and black points respectively, undetectable measures are represented as white symbols, and limits of detection are plotted. Error bars indicate means ± SEM. (* P < 0.05; ** P < 0.01; *** P < 0.001; **** P < 0.0001; ns, non-significant). For naïve donors, n=20 and for previously infected donors n=11.

**Figure S2:**
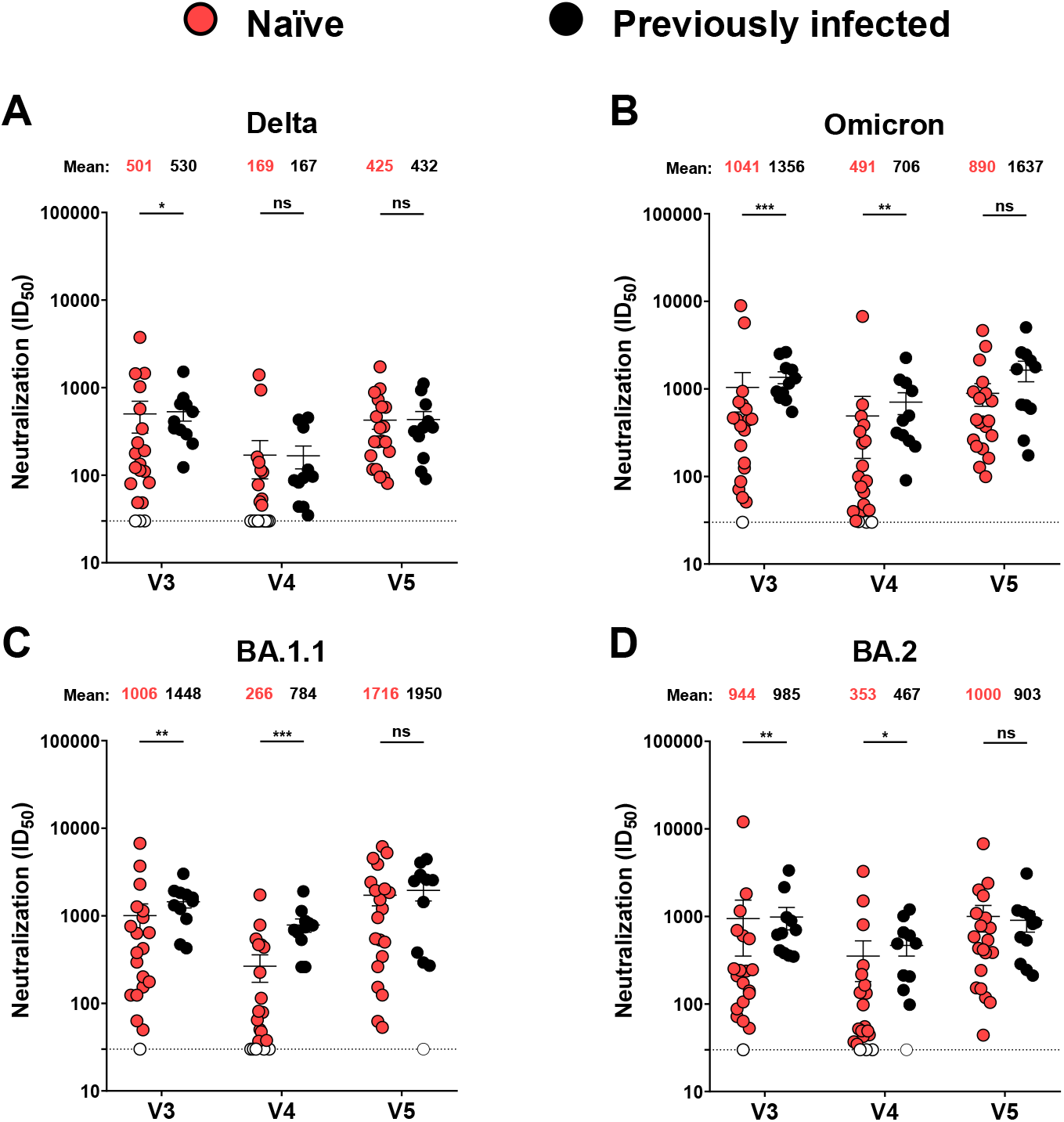
Neutralization activities in naive and previously-infected vaccinated individuals against several SARS-CoV-2 variants, Related to Figure 3. (**A-D**) Neutralizing activity was measured by incubating pseudoviruses bearing SARS-CoV-2 Delta S (**A**), Omicron S (**B**), BA.1.1 S (**C**) or BA.2 S (**D**) with serial dilutions of plasma for 1 h at 37°C before infecting 293T-ACE2 cells. Neutralization half maximal inhibitory serum dilution (ID50) values were determined using a normalized non-linear regression using GraphPad Prism software. Naive and PI donors are represented by red and black points, respectively. Plasma samples were grouped in different time points (V3, V4 and V5). Undetectable measures are represented as white symbols, and limits of detection are plotted. Error bars indicate means ± SEM (*p < 0.05; **p < 0.01; ***p < 0.001; ****p < 0.0001; ns, non-significant). For naive donors, n = 20 and for PI donors, n = 11.

**Figure S3:**
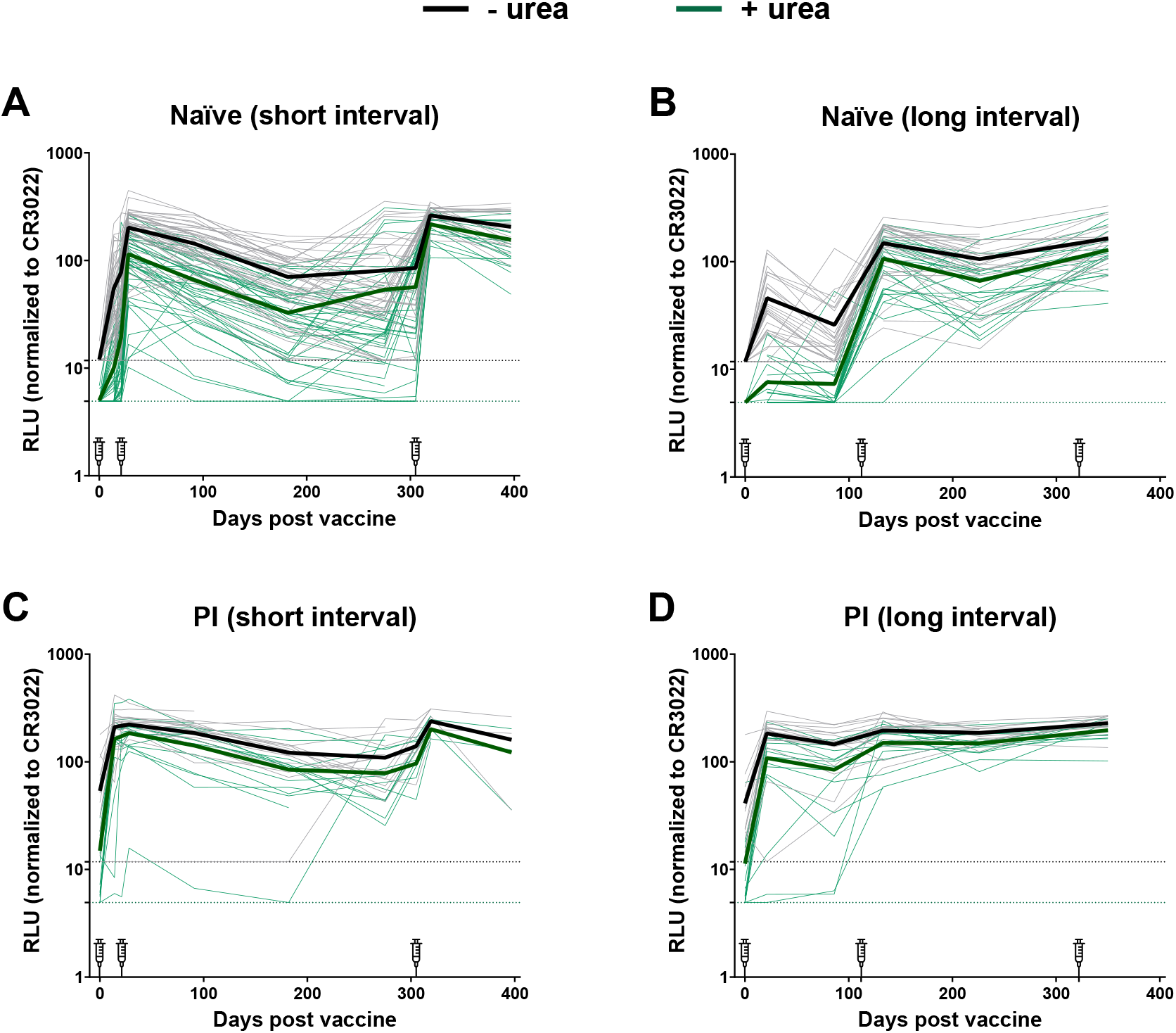
Comparison of the detection of RBD specific antibodies between ELISA and stringent ELISA in SARS-CoV-2 naïve and previously infected individuals vaccinated with a short or a long interval, Related to Figure 4. (**A-D**) Indirect ELISA was performed by incubating plasma samples from naïve (**A-B**) and PI (**C-D**) vaccinated donors after a short (**A, C**) or a long (**B, D**) interval with recombinant SARS-CoV-2 RBD protein. Anti-RBD Ab binding was detected using HRP-conjugated anti-human IgG. Relative light unit (RLU) values obtained were normalized to the signal obtained with the anti-RBD CR3022 mAb present in each plate. For ELISA (black curves), all the wash steps were made with washing buffer and for stringent ELISA (green curves), the wash steps were supplemented with 8M of urea. Each curve represents the normalized RLUs obtained with the plasma of one donor at every time point. Mean of each group is represented by a bold line. Limits of detection are plotted. For naïve donors n=46 for the short interval and n=30 for the long interval and for previously infected donors n=16 for the short interval and n=15 for the long interval.

**Figure S4:**
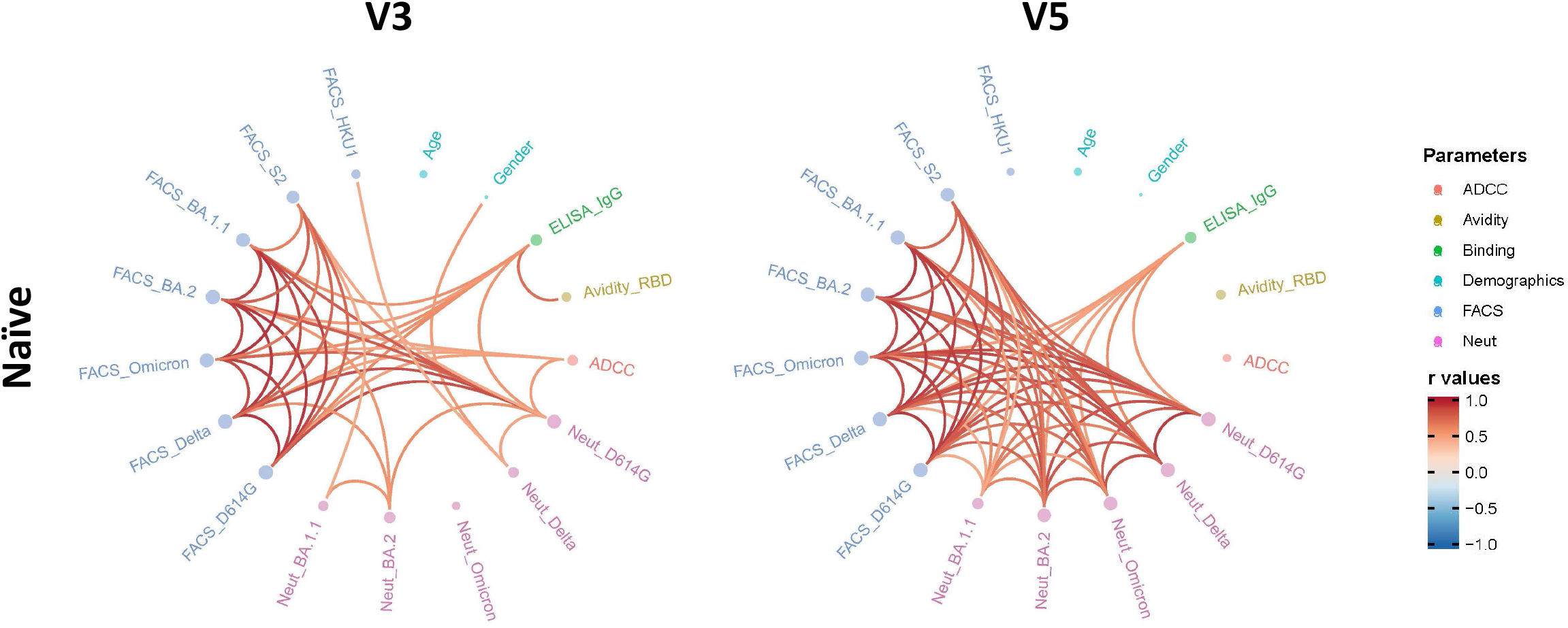
Mesh correlations of humoral response parameters after the second and the third dose of mRNA vaccine with the short interval regimen, Related to Figure 4. Edge bundling correlation plots where red and blue edges represent positive and negative correlations between connected parameters, respectively. Only significant correlations (p < 0.05, Spearman rank test) are displayed. Nodes are color coded based on the grouping of parameters according to the legend. Node size corresponds to the degree of relatedness of correlations. Edge bundling plots are shown for correlation analyses using two different datasets; i.e., SARS-CoV-2 naïve individuals vaccinated with the short interval at V3 and V5 respectively. n=20.

**Figure S5:**
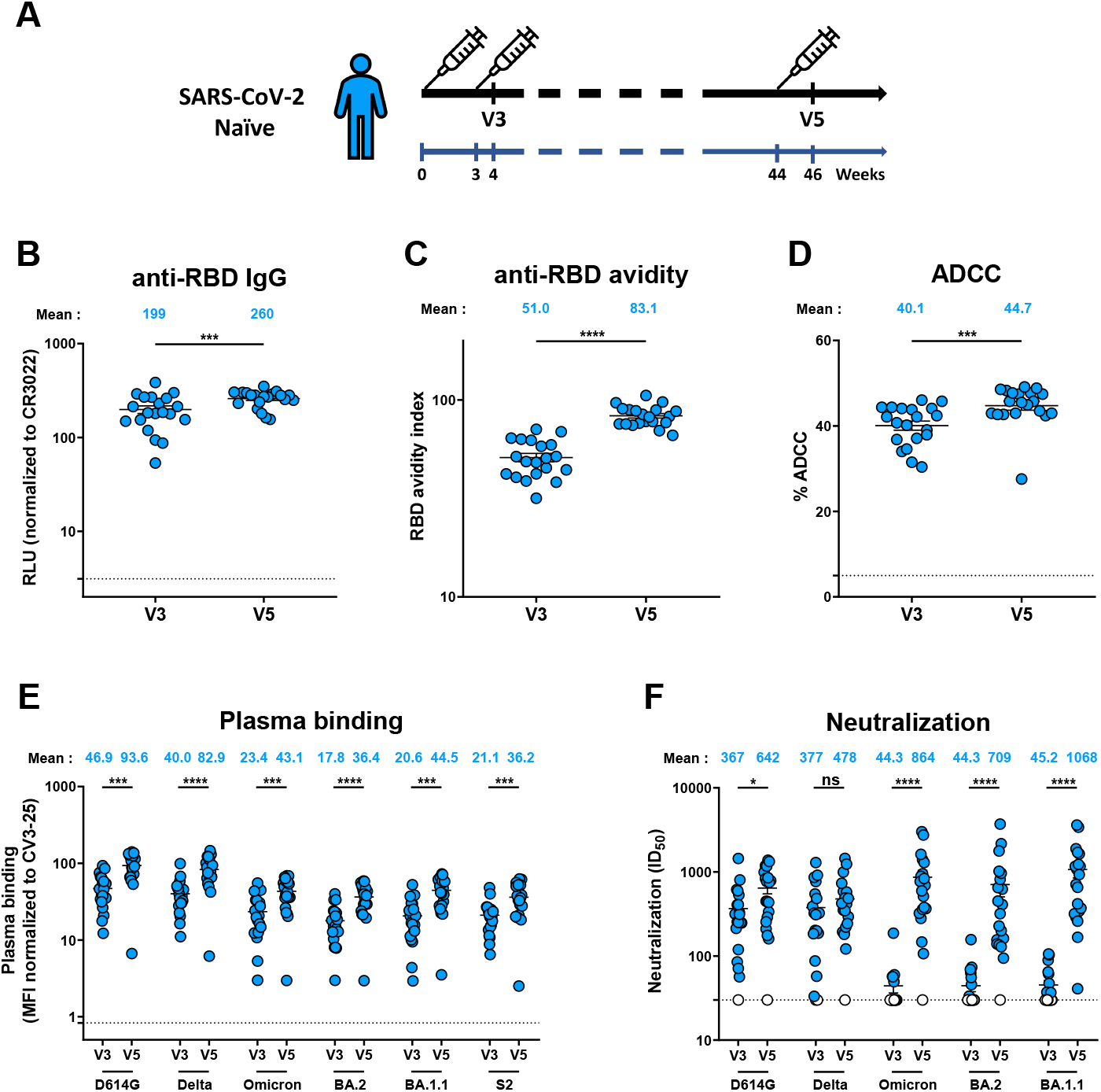
Humoral responses in SARS-CoV-2 naïve individuals that received a short dose interval, Related to Figure 4. (**A**) SARS-CoV-2 vaccine cohort design. (**B-F**) Humoral responses were measured in plasma samples collected after the second (V3) and the third dose (V5) from naïve donors that received a short dose interval. (**B**) Indirect ELISA was performed by incubating plasma samples with recombinant SARS-CoV-2 RBD protein. Anti-RBD Ab binding was detected using HRP-conjugated anti-human IgG. RLU values obtained were normalized to the signal obtained with the anti-RBD CR3022 mAb present in each plate. (**C**) Indirect ELISA and stringent ELISA was performed by incubating plasma samples with recombinant SARS-CoV-2 RBD protein. Anti-RBD Ab binding was detected using HRP-conjugated anti-human IgG. RBD avidity index corresponded to the value obtained with the stringent ELISA divided by that obtained with the ELISA. (**D**) CEM.NKr parental cells were mixed at a 1:1 ratio with CEM.NKr-S cells and were used as target cells. PBMCs from uninfected donors were used as effector cells in a FACS-based ADCC assay. (**E**) 293T cells were transfected with the indicated full-length S or the S2 subunit and stained with the CV3-25 Ab or with plasma and analyzed by flow cytometry. The values represent the MFI normalized by CV3-25 Ab binding. (**F**) Neutralizing activity was measured by incubating pseudoviruses bearing SARS-CoV-2 S glycoproteins with serial dilutions of plasma for 1 h at 37°C before infecting 293T-ACE2 cells. Neutralization half maximal inhibitory serum dilution (ID_50_) values were determined using a normalized non-linear regression using GraphPad Prism software. Undetectable measures are represented as white symbols, and limits of detection are plotted. Error bars indicate means ± SEM (*p < 0.05; **p < 0.01; ***p < 0.001; ****p < 0.0001; ns, non-significant). n = 20.

